# SCOPE: Integrating Organoid Screening and Clinical Variables Through Machine Learning for Cancer Trial Outcome Prediction

**DOI:** 10.64898/2026.04.10.26350512

**Authors:** Jean Bouteiller, Anna-Rose Gryspeert, Jérôme Caron, Lélia Polit, Gizem Altay, Mia Cabantous, Rafal Pietrzak, Fabiana Graziosi, Matéo Longarini, Kathryn Schutte, Jérôme Cartry, Jacques RR. Mathieu, Sabrina Bedja, Alice Boilève, Michel Ducreux, Diane-Laure Pagès, Fanny Jaulin, Gustave Ronteix

**Affiliations:** Orakl Oncology, Villejuif, France; INSERM U1279, Gustave Roussy, Université Paris-Saclay, Villejuif 94805, France; Département de Médecine Oncologique, Gustave Roussy, Université Paris-Saclay, Villejuif 94805, France; Institut Hospitalo-Universitaire PRISM, Gustave Roussy, Université Paris-Saclay, Villejuif 94805, France

**Keywords:** patient-derived organoids, clinical trial prediction, colorectal cancer, pancreatic cancer, drug development, precision oncology

## Abstract

**Background:** Predicting whether a treatment will demonstrate meaningful clinical benefit before committing to a large-scale trial remains a major unmet need in oncology. Patient-derived organoids (PDOs) recapitulate individual tumor drug sensitivity, but have not been used to forecast population-level trial outcomes. We developed SCOPE (Screening-to-Clinical Outcome Prediction Engine), a platform that integrates PDO drug screening with clinical prognostic modeling to predict arm-level median progression-free survival (mPFS) and objective response rate (ORR) without access to any trial outcome data.

**Patients and methods:** SCOPE was trained on 54 treatment lines from 52 independent patients with metastatic colorectal cancer (mCRC, *n*=15) and metastatic pancreatic ductal adenocarcinoma (mPDAC, *n*=39) with matched clinical data and PDO drug screening across 9 compounds. A Clinical Score module captures baseline prognosis; a Drug Screen Score module quantifies treatment-specific organoid sensitivity. To predict trial outcomes, synthetic patient profiles are generated from published eligibility criteria and matched to a biobank of 81 PDO lines. Predictions were externally validated against 32 arms from 23 published trials, treatment ranking was assessed across 8 head-to-head comparisons, and prospective applicability was tested for Daraxonrasib (RMC-6236), a novel pan-RAS inhibitor in mPDAC.

**Results:** Predicted mPFS strongly agreed with published outcomes (*R*^2^=0.85, MAE=0.82 months; Pearson *r*=0.92, *P* <0.001), approaching the empirical concordance between two independently measured clinical endpoints (ORR vs. mPFS, *R*^2^=0.87). ORR prediction was similarly robust (*R*^2^=0.71, MAE=7.3 percentage points). Integrating organoid and clinical data significantly out-performed either alone (*P* =0.001). SCOPE correctly identified the superior arm in 7 of 8 head-to-head comparisons (88%, *P* <0.05). Applied to Daraxonrasib prior to phase 3 data availability, the platform predicted superiority over standard chemotherapy in KRAS-mutant mPDAC, consistent with emerging trial data.

**Conclusion:** By combining functional organoid drug screening with clinical modeling, SCOPE generates calibrated efficacy predictions for both established regimens and novel agents without prior trial data. This approach could support clinical trial design, treatment arm selection, and go/no-go decisions, offering a new tool to improve the efficiency of gastrointestinal cancer drug development.

## INTRODUCTION

Oncology drug development is marked by an exceptionally high failure rate, with only 3–5% of clinical programs achieving regulatory approval (1). Lack of clinical efficacy accounts for 40–50% of these failures (2; 3), underscoring a fundamental challenge: the inability to reliably predict whether a given treatment will produce a meaningful clinical benefit in the intended patient population before committing to the cost, duration and patient impact of a largescale trial. Tools that can anticipate treatment efficacy earlier in the development cycle would represent a major advance for the field.

Computational approaches to predict clinical trial outcomes have gained traction in recent years (4; 5) and demonstrate the potential of machine learning for trial-level prediction. However, these approaches are fundamentally dependent on historical trial data (drug–disease associations, prior clinical outcomes, and trial metadata), making them difficult to apply to novel therapeutic agents for which no prior clinical experience exists. Moreover, they predict binary success or failure rather than quantitative efficacy endpoints such as median progression-free survival (mPFS) or objective response rate (ORR), limiting their utility for trial design optimization and comparative effectiveness assessment.

In parallel, patient-derived organoids (PDO) have emerged as the most clinically relevant in vitro model for drug sensitivity assessment. Unlike traditional cell lines, PDOs retain the histological architecture, genetic landscape, and intratumoral heterogeneity of the original tumor, and landmark studies have demonstrated that ex-vivo PDO drug responses recapitulate individual patient outcomes across gastrointestinal cancers (6; 7), particularly for chemotherapy, where genetic biomarkers remain poorly predictive (8; 9; 10; 11). Meanwhile, artificial intelligence approaches that integrate multimodal clinical data, including imaging, genomics, clinical records, and molecular profiling, have shown impressive performance in predicting individual patient prognoses (12; 13). However, both paradigms operate at the individual patient level and have not been extended to simulate clinical trial outcomes across a treated population.

Here, we present SCOPE (Screening-to-Clinical Outcome Prediction Engine), an integrated computational platform that bridges this gap by combining clinical prognostic scoring with PDO drug screening to predict quantitative, population-level treatment outcomes. Applied to a biobank of 81 organoid lines from patients with metastatic colorectal cancer (mCRC) and metastatic pancreatic ductal adenocarcinoma (mPDAC) screened against 9 compounds, SCOPE generates arm-level predictions of mPFS and ORR without access to any trial outcome data. To our knowledge, this is the first study to use PDO drug screening to predict population-level clinical trial outcomes.

## METHODS

### Study design and overview

This study is a retrospective validation of SCOPE, Orakl Oncology’s predictive platform using published clinical trial outcomes as external benchmarks. We evaluated SCOPE’s predictive performance through three complementary analyses of increasing clinical relevance: (1) calibration of predicted versus observed treatment outcomes across individual trial arms; (2) discrimination accuracy in identifying the superior treatment arm in head-to-head comparisons; (3) pseudo-prospective prediction for Daraxonrasib (RMC-6236), a novel therapeutic agent in an ongoing clinical trial.

SCOPE was developed on the development cohort, then applied prospectively to simulate trial outcomes without access to published results. No patient-level data from validation trials was used; all comparisons are against published aggregate endpoints.

### Patient data

All patient data and tumor specimens used in this study were collected under institutional review board–approved clinical protocols at Gustave Roussy hospital as part of STING (NCT04932525), ORGANOTREAT (NCT05267912) and MATCH-R (NCT02517892) clinical trials. All patients provided written informed consent for tumor sampling, organoid generation, and the use of clinical data for research purposes.

### Development cohort

The development cohort was sampled from the Gustave Roussy cohort to which we applied several selection criteria, meant to facilitate model training. We selected patients having received a metastatic treatment line following PDO sampling and we excluded patients that did not have a recorded progression date for the corresponding treatment line at the time of collection, and patients that interrupted the treatment due to causes other than tumor progression (including, but not limited to, toxicity, loss of contact, etc.). As a result, the SCOPE model was developed on 54 treatment lines from a cohort of 52 patients (15 mCRC and 37 mPDAC) treated at Gustave Roussy. Comprehensive clinical and molecular data were available for all patients, including demographics, ECOG performance status, molecular profiling, treatment history, and outcomes. This was a heavily pretreated population (more than half of patients at least in third line) with a mPFS of 2 months (IQR: 1.6–2.6) and a response rate of 7.4%. Tumor tissues were obtained from core needle biopsies, ascites or pleural effusions. Detailed baseline characteristics can be found in Supplementary Table 1.

### PDO biobank and drug screening

#### Patient-Derived Organoid Lines

Established patient-derived organoid (PDO) lines from patients with mCRC and mPDAC were obtained from the INSERM U1279 laboratory at Gustave Roussy (Villejuif, France). PDO lines were generated from fresh tumor specimens (surgical resection or core needle biopsies) following previously described protocols for tissue processing, tumor digestion, and tumor cell isolation (10; 11). The cohort of 78 patients from which the organoid cohort was derived and the biobank of 81 PDOs with drug screening information are described in Supplementary Table 2.

#### Organoid Culture

PDOs were cultured in Cultrex UltiMatrix Reduced Growth Factor Basement Membrane Extract (Bio-Techne; BME001-05) and passaged every 7–12 days. At confluence, organoids were dissociated by incubation with TrypLE Express 1X (Thermo Fisher Scientific; 12605010) at 37°C with intermittent mixing every 5 minutes until single cells or small clusters were obtained. Dissociation was stopped by addition of DMEM GlutaMAX (Life Technologies; 31966047) supplemented with 10% fetal bovine serum (FBS; Life Technologies; A5670801) and 1% penicillin–streptomycin (Gibco; 15140122). Cells were pelleted by centrifugation, washed once with the same supplemented medium, and repelleted. The cell pellet was resuspended in Cultrex UltiMatrix at a density of 500 cells/µL, plated as domes, and incubated for 20 minutes at 37°C to allow matrix polymerization. PDO culture medium modified from Fujii et al. (14) supplemented with IntestiCult™ Organoid Growth Medium (StemCell™ Technologies; 06010) and 10 µM Y-27632 ROCK inhibitor (Bio-Techne; 1254/10) was then added, and plates were maintained at 37°C in a humidified atmosphere with 5% CO_2_.

#### Drug Screening Assay

Fully grown organoids were dissociated using TrypLE Express 1X (Thermo Fisher Scientific; 12605010) and filtered to obtain a singlecell suspension. Cells were pelleted, resuspended in culture medium containing 2% Matrigel (Corning; 356231) and 10 µM Y-27632 (Bio-Techne; 1254/10), and dispensed into low-adhesion 384-well microplates (Revvity; 6057802) at a density of 1,500 cells per well in 50 µL. Plates were incubated at 37°C with 5% CO_2_ for two days to allow organoid formation. On day 2 post-seeding, 25 µL of fresh culture medium containing drugs at the indicated concentrations was added to each well.

On day 7 post-seeding (five days of drug exposure), cell viability was assessed using the CellTiter-Glo luminescent cell viability assay (Promega; G7570). Briefly, 25 µL of CellTiter-Glo reagent was added to each well, plates were shaken for 5 minutes on an orbital shaker and incubated at room temperature for 20 minutes. Luminescence was measured using a Synergy LX multi-mode microplate reader (Agilent BioTek).

#### Drug Response Analysis

For each drug, mean luminescence values at each concentration were normalized to the corresponding solvent control (vehicle-treated) wells to calculate percentage cell viability. Dose–response curves were generated by plotting percentage viability as a function of drug concentration. A four-parameter sigmoidal (Hill equation) model was fitted to each dose–response curve to derive half-maximal effective concentration (EC_50_) and half-maximal inhibitory concentration (IC_50_) values.

#### Drug Panel

A panel of 9 compounds, comprising both cytotoxic chemotherapies and targeted agents, was selected for PDO screening either as single agents (6) or combinations (4). All compounds were sourced from MedChemExpress unless otherwise noted:

- **Cytotoxic chemotherapies (***n***=6):** 5-fluorouracil (HY-90006), SN-38 (active metabolite of irinotecan; HY-13704), oxaliplatin (HY-17371), paclitaxel (HY-B0015), trifluridine/tipiracil (HY-16478), gemcitabine (HY-17026)
- **Targeted therapies (***n***=1):** RMC-6236 (pan-RAS inhibitor; HY-148439)
- **Antibody–drug conjugates (***n***=2):** tusamitamab ravtansine (HY-P99542), sacituzumab govitecan (HY-132254)

#### Quality Control

Organoid cultures were routinely tested for Mycoplasma contamination. Prior to passaging, 500 µL of culture medium supernatant was collected, heated at 95°C for 10 minutes, and centrifuged at 13,000 rpm to pellet cellular debris. Supernatant was submitted to Eurofins Genomics for quantitative PCR-based Mycoplasma detection.

All drug concentrations were screened in at least triplicate, and statistical outlier detection was applied to replicate measurements before averaging. Luminescence values were normalized using an outlier-robust method based on the 95th percentile of vehicle-treated wells on each plate, rather than the plate maximum, to mitigate the influence of aberrant control wells. Experiments flagged for general technical failures (e.g., culture medium issues, robotic dispensing errors) were excluded from analysis. Individual data points identified as experimental artifacts were removed prior to curve fitting using an outlier detection framework. Dose–response curves with residuals exceeding a predefined noise threshold were excluded to ensure reliable sigmoid fits.

### SCOPE platform architecture

SCOPE is an AI platform that integrates patient clinical data and PDO drug screening results to generate outcome predictions, such as patients’ PFS and probability of response to treatment.

The prediction pipeline, presented in Figure 1A, consists of three core modules that generate patient predictions.

**Figure 1:**
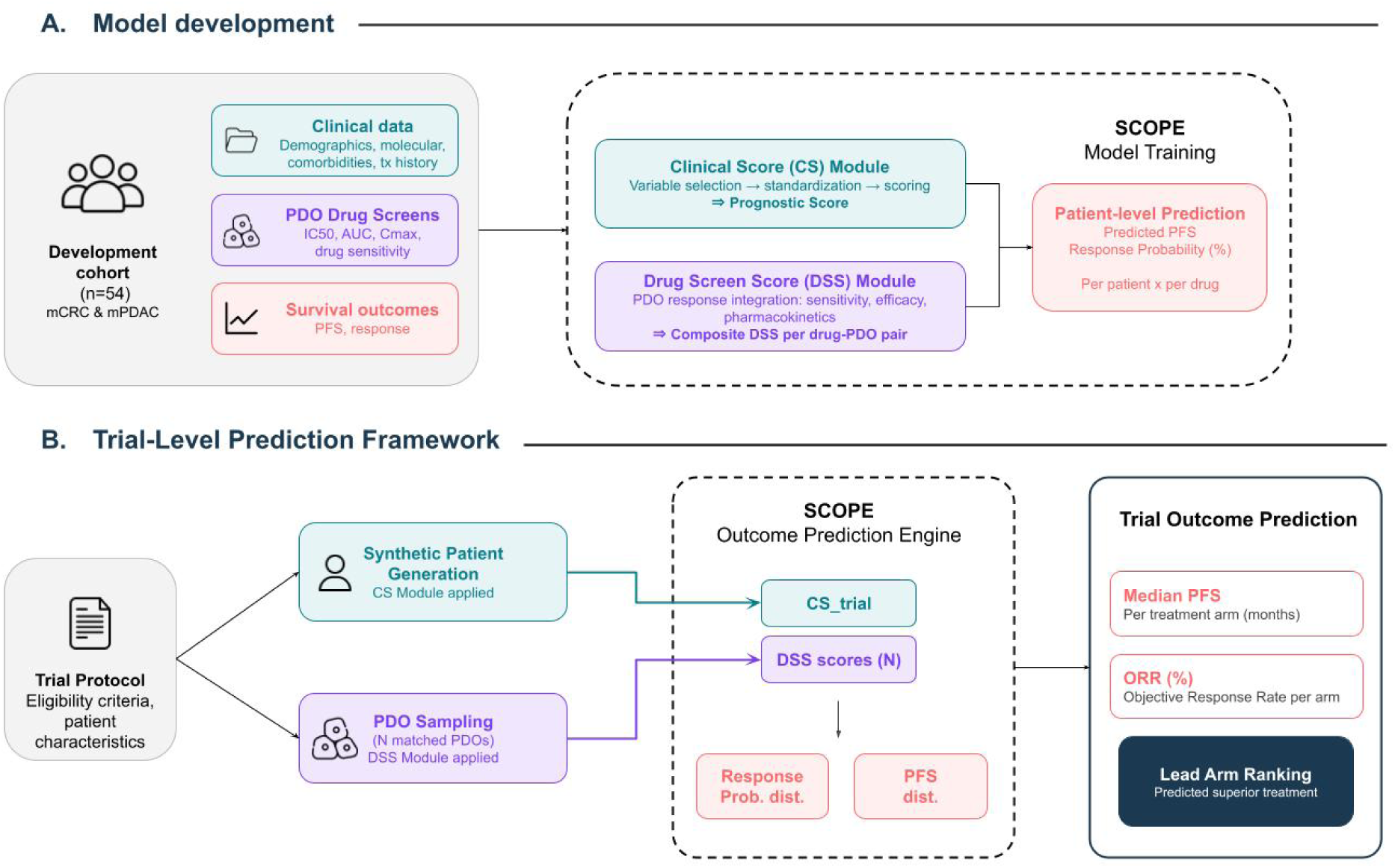
Overview of the SCOPE platform architecture and trial-level prediction framework. **(A)** Model development. SCOPE was trained on a cohort of 54 treatment lines of mCRC and mPDAC from Gustave Roussy, with matched patient data, patient-derived organoid drug screening results, and survival outcomes. The platform comprises two scoring modules: a Clinical Score (CS) module that integrates demographic, molecular, and treatment history variables into a composite prognostic score and a Drug Screen Score (DSS) module that quantifies treatment-specific response potential from PDO sensitivity assays incorporating drug sensitivity, efficacy, and other drug parameters. The outcome prediction model integrates CS and DSS to generate patient-level predictions of PFS and treatment response probability. **(B)** Trial-level prediction framework. To simulate clinical trial outcomes, SCOPE follows a four-stage process: First, a synthetic patient profile is generated from trial eligibility criteria and published baseline characteristics and scored using the CS module. Second, PDOs matching the trial cancer type and molecular criteria are sampled from the biobank, and their DSS scores are retrieved for each treatment arm. Then, SCOPE generates PFS and objective response distributions from the combined CS and DSS inputs. Finally, predicted mPFS and ORR are computed for each treatment arm, and the predicted superior arm is compared against published trial results.

#### Clinical Score (CS) module

The clinical score module quantifies patient baseline prognosis independent of treatment selection, integrating comprehensive clinical and molecular data: demographics, comorbidities, disease characteristics, molecular and biomarker profiles, and treatment history.

The CS module employs a multi-stage machine learning pipeline: first, it extracts clinical data from patient records, then, it automatically identifies the relevant prognostic features using published literature and clinical guidelines and finally, it integrates the selected variables into a continuous score where a higher value indicates a better prognosis. Detailed CS methodology and comprehensive validation will be reported in a separate publication focused on machine learning approaches for clinical risk prediction.

#### Drug Screen Score (DSS) module

The DSS is a proprietary score that integrates multiple PDO drug response metrics including measures of drug sensitivity (EC_50_), maximal efficacy, and pharmacological properties to generate a composite score for each PDO–drug pair, where higher values indicate greater treatment response. Detailed DSS methodology including the proprietary integration algorithm and comprehensive PDO–clinical correlation studies will be reported in a dedicated publication.

#### Outcome prediction module

SCOPE integrates the CS and DSS in a linear model to predict individual patient PFS and treatment response probability. Model parameters were selected using 5-fold cross-validation, and the final coefficients were estimated by fitting an Elastic Net–penalized linear model to the full development cohort. This study focuses on external validation of trial-level predictions. External validation of individual patient-level predictions is ongoing in an independent cohort and will be reported separately.

### Trial-level prediction framework

We used SCOPE to predict population-level clinical trial outcomes as presented in Figure 1B.

First, trial eligibility criteria and published baseline characteristics (molecular subgroup, treatment line, ECOG performance status, median age) are used to generate a representative patient profile, from which a trial-level Clinical Score (CS_trial_) is obtained using the CS module described above. Next, all PDOs matching the trial’s cancer type and the molecular subgroup criteria are selected from the biobank and screened against the trial-specific drug regimen to obtain individual Drug Screen Scores. SCOPE then combines the CS_trial_ score with each PDO’s DSS to generate patient-level PFS and treatment response predictions. Finally, population-level end-points are derived by computing the median PFS and objective response rate across the set of individual predictions.

### Statistical analysis

Statistical analyses were designed to evaluate the agreement between SCOPE’s simulated trial outcomes and observed clinical trial results at both the arm level and the treatment-comparison level.

#### Calibration

For each trial arm, we compared predicted mPFS and objective response rate (ORR) against published values. Agreement was quantified using Pearson’s correlation coefficient (*r*), Spearman’s rank correlation (*ρ*), the coefficient of determination (*R*^2^), mean absolute error (MAE), and root mean square error (RMSE). These metrics were computed across all arms pooled and stratified by cancer type (mCRC and mPDAC). To characterize the clinical relevance of prediction errors, we reported the proportion of arms for which predicted values fell within *±*1, *±*2, and *±*3 months of observed mPFS, and within *±*10, *±*20, and *±*30 percentage points of observed ORR.

#### Treatment ranking

For each head-to-head comparison of treatment arms, concordance was defined as the correct identification of the trial arm with higher observed mPFS. The overall concordance rate was tested against the null hypothesis of random performance (50%) using a one-sided exact binomial test. Confidence intervals for the concordance rate were computed using the Clopper–Pearson exact method. Concordance was also evaluated on the subset of comparisons restricted to trials with officially reported superiority of one arm over the other.

#### Model component analysis

To evaluate the relative contribution of the CS and DSS to prediction accuracy, we compared the full SCOPE model against each scoring module in isolation. For each trial arm, the median CS and median DSS across all matched PDOs were computed and used as single-score predictors of observed mPFS and ORR. Agreement was quantified using *R*^2^ for each predictor, and uncertainty was estimated using a nonparametric bootstrap procedure: trial arms were resampled with replacement (1,000 iterations), and *R*^2^ was computed for each resample. The 95% confidence interval was derived from the 2.5th and 97.5th percentiles of the bootstrap distribution. To test whether the full model outperformed the CS alone or DSS alone, we computed *R*^2^ for both predictors on each of 1,000 bootstrap resamples of trial arms. The *p*-value was defined as the proportion of resamples in which 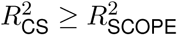 and 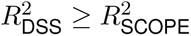

#### Benchmark analysis

As an external benchmark, we compared the correlation between predicted and observed mPFS against the correlation between observed ORR and observed mPFS. This benchmark reflects the empirical agreement between two established clinical endpoints and provides an upper bound for the accuracy that can be expected from any predictive model operating without access to patient-level survival data.

#### Subgroup analyses

To explore potential sources of heterogeneity in prediction accuracy, we compared absolute arm-level prediction errors across subgroups defined by trial publication era, treatment line, treatment regimen class, and organoid panel size, using Mann–Whitney U tests.

All statistical tests were two-sided unless otherwise specified, with significance set at *α* = 0.05. Analyses were conducted in Python 3.13.5 using SciPy 1.17.0 and lifelines 0.30.0.

## RESULTS

### Result 1: Calibration of predicted treatment outcomes

#### Data

To evaluate SCOPE’s ability to predict population-level treatment outcomes, we assembled a dataset of published clinical trials in mCRC and mPDAC in which patients were treated with drugs available in the PDO biobank screening panel, and for which median PFS and ORR were reported per arm. The full validation dataset comprised 32 treatment arms from 23 trials described in Supplementary Table 4 and summarized in Table 1. On average, 26 PDOs are used per trial arm, ranging from 3 to 40 with a median of 25. Observed median PFS was available for 31 out of 32 arms in the calibration set, and observed ORR was available for all arms.

**Table 1:** Characteristics of the validation dataset.

| **Characteristic** | **N (%)** |
| --- | --- |
| Total arms | 32 |
| Total trials | 23 |
| Year, median (range) | 2006 (2000–2024) |
| **Disease** |  |
| CRC | 15 (46.9%) |
| PDAC | 17 (53.1%) |
| **Phase** |  |
| Phase 1 | 2 (6.2%) |
| Phase 1–2 | 2 (6.2%) |
| Phase 2 | 4 (12.5%) |
| Phase 2–3 | 2 (6.2%) |
| Phase 3 | 22 (68.8%) |
| **Setting** |  |
| Metastatic | 32 (100.0%) |
| **Treatment line** |  |
| 1L | 21 (65.6%) |
| 2L | 5 (15.6%) |
| 2L or beyond | 4 (12.5%) |
| 3L or beyond | 1 (3.1%) |
| **Molecular subgroups** |  |
| EGFR Expressing | 1 (3.1%) |
| KRAS Mutant | 2 (6.2%) |
| KRAS WT | 1 (3.1%) |
| KRAS Mutant G12X | 1 (3.1%) |
| **Treatment type** |  |
| Antibody–drug conjugate | 3 (9.4%) |
| Chemo | 28 (87.5%) |
| Targeted therapy | 1 (3.1%) |

CRC, colorectal cancer; PDAC, pancreatic ductal adenocarcinoma; 1L, first-line; 2L, second-line; 3L, third-line; EGFR, epidermal growth factor receptor; KRAS, Kirsten rat sarcoma viral oncogene homolog; RAS, rat sarcoma; WT, wild-type.

#### Median PFS prediction

Across the 31 evaluable arms in the calibration set, SCOPE’s predicted mPFS values ranged from 0.69 to 10.11 months (median: 5.12 months). Predicted mPFS showed strong agreement with observed values (*R*^2^ = 0.85, MAE = 0.82 months; Figure 2, Supplementary Table 3A). All correlation coefficients were highly significant (Pearson *r* = 0.92, Spearman *ρ* = 0.91; both *P ≤*0.001).

**Figure 2:**
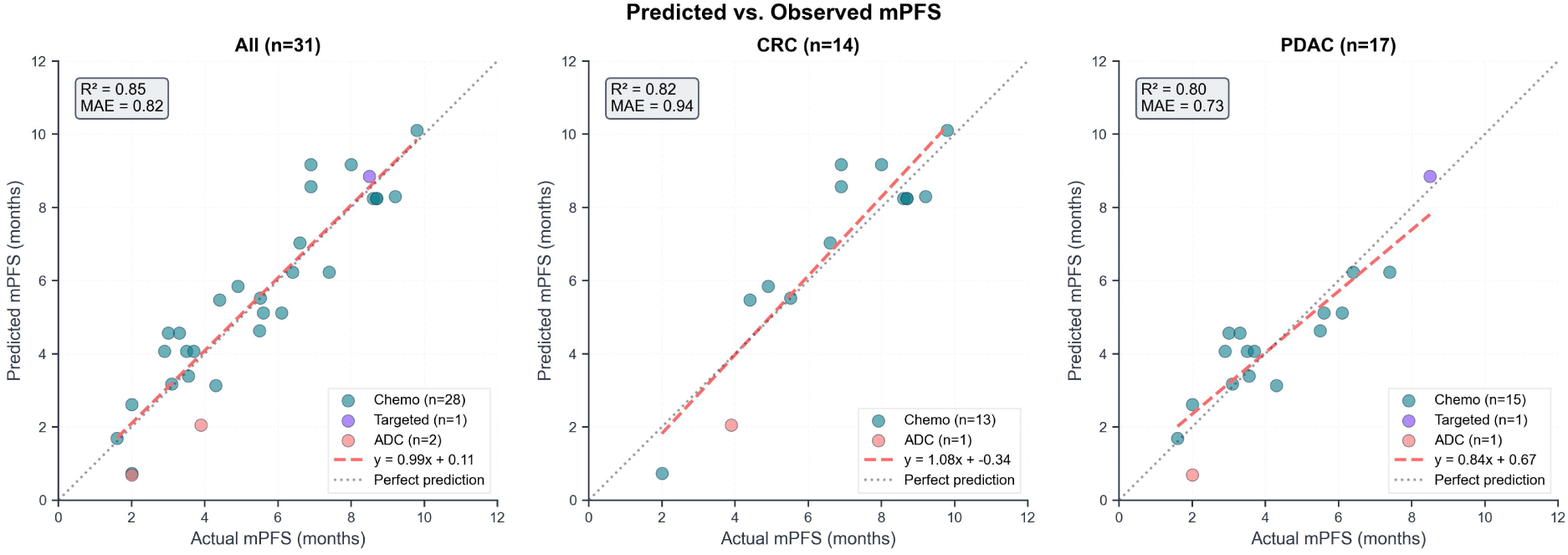
Predicted versus observed median PFS across clinical trial arms. Predicted mPFS is plotted against published mPFS for all trial arms (left; *n*=31), CRC trials (center; *n*=14), and PDAC trials (right; *n*=17). Drug regimen is color-coded to identify chemotherapy regimens, targeted therapy regimens (as single therapy or in combination with chemotherapy) and antibody–drug conjugates (ADC).

Calibration was similar for CRC (*R*^2^ = 0.82, MAE = 0.94 months) and PDAC (*R*^2^ = 0.80, MAE = 0.73 months). Overall, 97% of predicted mPFS values fell within *±*2 months of the observed value and all within *±*3 months.

Regression slope lower than 1 for PDAC indicated moderate compression of predictions toward the mean: the model tended to overestimate mPFS for arms with short observed survival and underestimate it for arms with long observed survival. Despite this compression in absolute values, the model preserved strong rank ordering across arms (Spearman *ρ* = 0.91), suggesting that the attenuation affects the scale of predictions more than their relative accuracy.

#### ORR prediction

Across 32 arms, SCOPE’s predicted ORR showed strong agreement with observed values (*R*^2^ = 0.71, MAE = 7%; Figure 3, Supplementary Table 3B). All correlations were highly significant (Pearson *r* = 0.84, Spearman *ρ* = 0.86; both *P* < 0.001). Overall, 97% of predictions fell within *±*20 percentage points of the observed ORR, and all within *±*30 percentage points.

**Figure 3:**
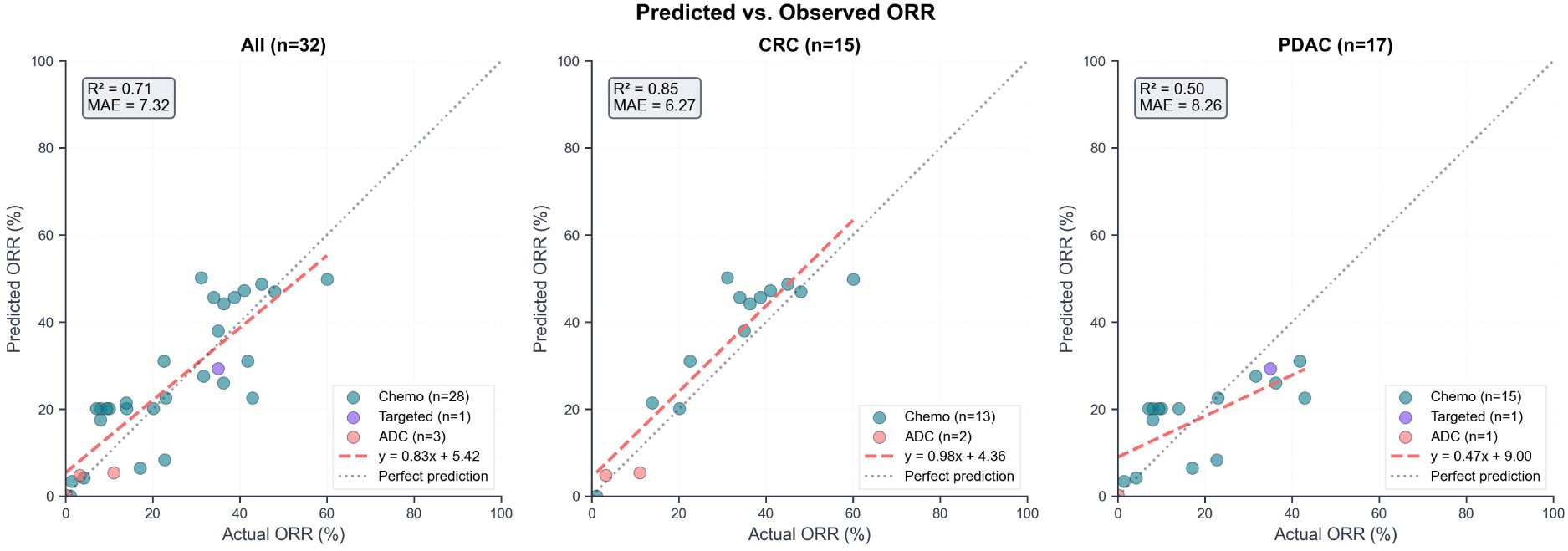
Predicted versus observed objective response rates across clinical trial arms. Predicted ORR is plotted against published ORR for all trial arms (left; *n*=32), CRC trials (center; *n*=15), and PDAC trials (right; *n*=17). Drug regimen is color-coded to identify chemotherapy regimens, targeted therapy regimens (as single therapy or combination with chemo), and antibody–drug conjugates (ADC).

ORR calibration was stronger in CRC (*R*^2^ = 0.85, MAE = 6%) than in PDAC (*R*^2^ = 0.50, MAE = 8%). In PDAC, the lower *R*^2^ was accompanied by a preserved rank correlation (Spearman *ρ* = 0.82, *P* < 0.001), indicating that the model correctly ordered arms by response rate despite weaker absolute precision.

As with mPFS, the regression slope below 1.0 indicated a slight compression toward the mean, with the model overestimating ORR for poorly responding arms and underestimating it for highly responsive ones.

#### Performance benchmark

To contextualize the model’s calibration performance, we compared the agreement between predicted and observed mPFS (*R*^2^ = 0.85) against the agreement between two independently observed clinical endpoints, ORR and mPFS, across the same 31 trial arms. Observed ORR and observed mPFS were strongly correlated (*R*^2^ = 0.87), indicating high arm-level concordance between these endpoints in our validation dataset. The model’s predicted mPFS achieved approximately 98% of this inter-endpoint agreement, suggesting that organoid-informed predictions match the level of concordance seen between independently measured clinical outcomes, without requiring trial execution.

#### Model robustness

To assess whether prediction accuracy varied systematically with trial or model characteristics, we compared absolute mPFS prediction errors across four stratifications (Figure 4). Prediction error did not differ significantly between subgroups, indicating that model performance was stable across trial era, clinical setting, treatment class, and organoid panel size.

**Figure 4:**
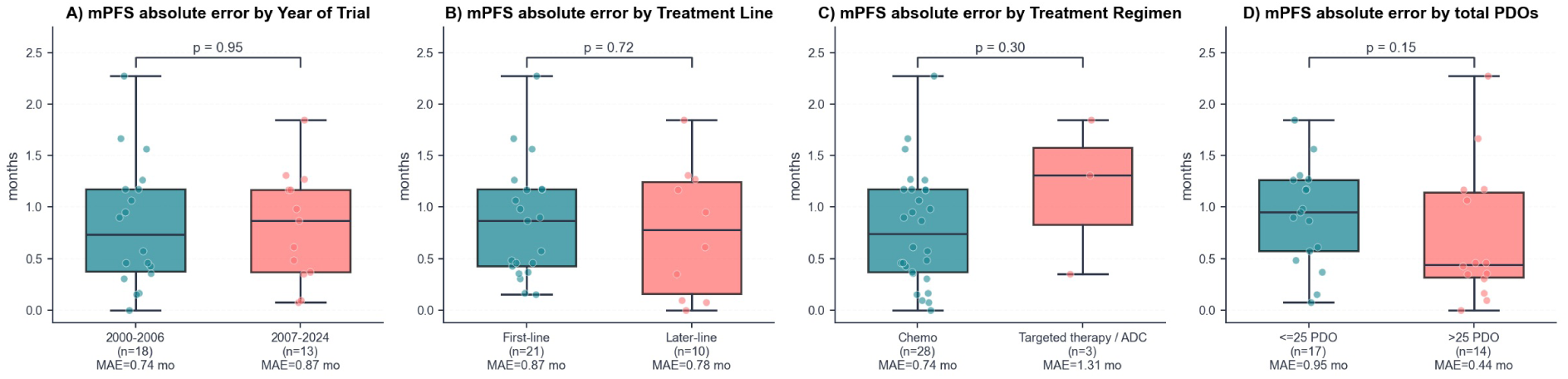
Prediction error is consistent across trial characteristics. Absolute error between predicted and observed median PFS is shown stratified by trial publication year (A; before vs. after median year of 2006), treatment line (B; first-line vs. later-line), treatment regimen (C; chemotherapy vs. targeted therapy and ADC), and organoid panel size (D; fewer or more than the median of 25 PDOs). Subgroup size and median MAE values are displayed beneath each subgroup. Boxes represent interquartile ranges with median lines; whiskers extend to 1.5*×* IQR; individual data points are overlaid.

#### Contribution of model components

To assess whether the integration of clinical and drug screening data contributes independently to prediction accuracy, we compared the full model against each module in isolation. For each trial arm, we computed the median CS and median DSS across the organoid panel and evaluated the concordance of each with observed mPFS and ORR, as shown in Figure 5.

**Figure 5:**
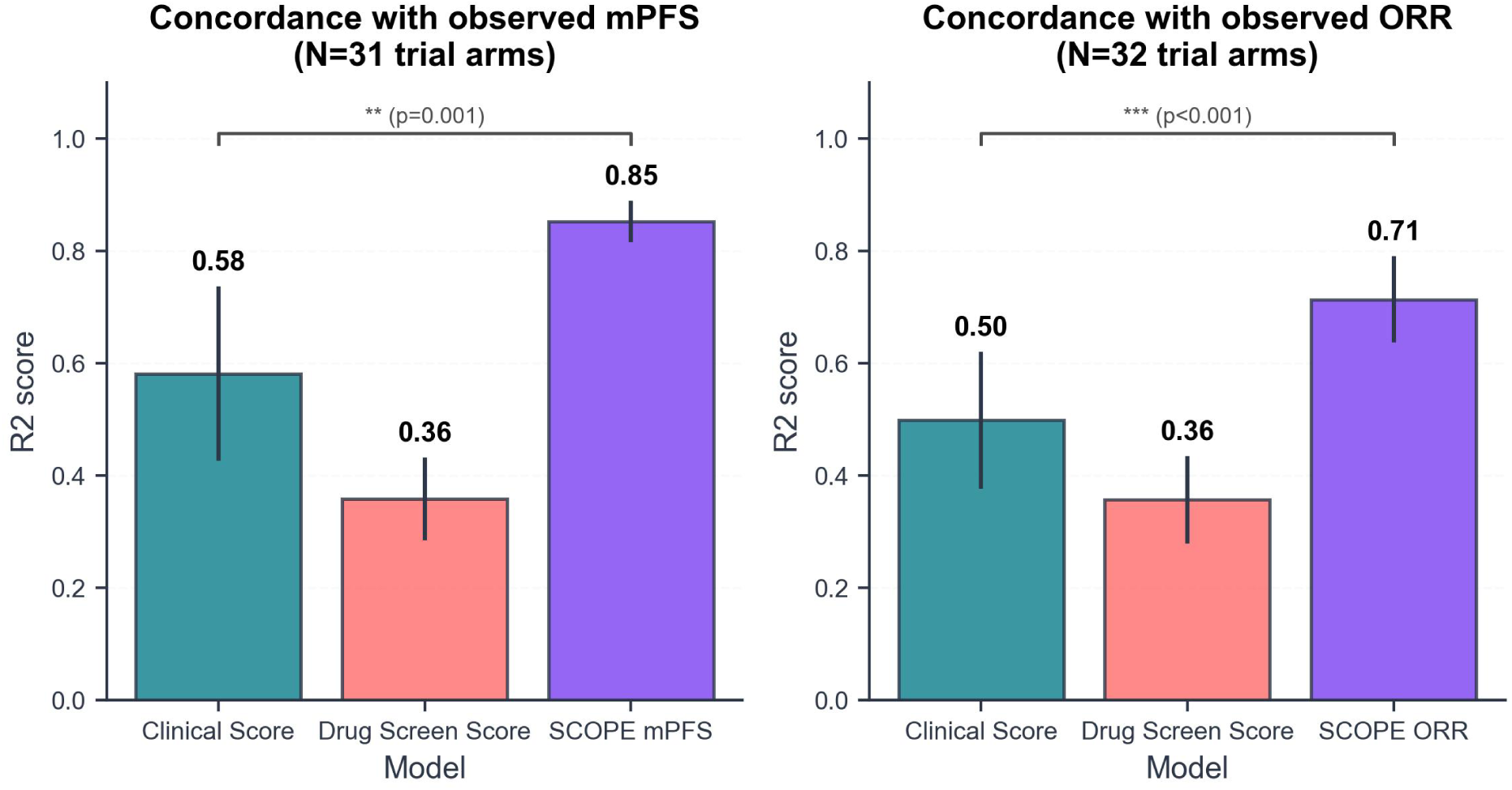
Contribution of individual model components to prediction accuracy. *R*^2^ between predicted and observed values is shown for the clinical score (CS) alone, the drug screen score (DSS) alone, and the full SCOPE model, evaluated against observed median PFS (left; *N* = 31 arms) and observed ORR (right; *N* = 32 arms). For each trial arm, the median CS and median DSS across the organoid panel were used as single-score predictors. Error bars represent the standard deviation of *R*^2^ estimates obtained from 1,000 bootstrap resamples of trial arms.

For mPFS, the median CS alone achieved moderate concordance with observed values (*R*^2^ = 0.58, 95% CI: 0.29–0.84), while the median DSS alone showed weaker agreement (*R*^2^ = 0.36, 95% CI: 0.23–0.54). The full model’s predicted mPFS achieved the highest point estimate (*R*^2^ = 0.85, 95% CI: 0.78–0.92), significantly outperforming the DSS alone and the CS alone (paired bootstrap *P* = 0.001). A similar pattern was observed for ORR: the median CS explained a larger share of variance (*R*^2^ = 0.50, 95% CI: 0.24–0.71) than the median DSS (*R*^2^ = 0.36, 95% CI: 0.22–0.55), while the full model again achieved the highest concordance (*R*^2^ = 0.71, 95% CI: 0.55–0.85) significantly outperforming the CS alone (*P* < 0.001). Across both endpoints, the integrated model consistently yielded numerically higher concordance than either component alone. Notably, the CS alone cannot distinguish between treatment arms within a given trial, as it produces identical predictions for all arms sharing the same patient population, a limitation addressed in the subsequent treatment ranking analysis.

Having established that the integrated model provides calibrated predictions that depend on both clinical and drug screening inputs, we next assessed whether this calibration translates to correct treatment ranking in head-to-head comparisons.

### Result 2: Treatment arm ranking prediction

#### Data

To assess treatment ranking performance, we derived a head-to-head comparison subset by selecting trials in which both arms were evaluable. The characteristics of the 8 pairwise comparisons from 16 arms used for treatment ranking validation are summarized in Table 2, and the full list of pairwise comparisons along with predictions is shown in Supplementary Table 5.

**Table 2:** Characteristics of the treatment ranking validation dataset.

| Characteristic | N (%) |
| --- | --- |
| Total arms | 16 |
| Total trials | 8 |
| Year, median (range) | 2008 (2000–2020) |
| **Disease** |  |
| CRC | 6 (37.5%) |
| PDAC | 10 (62.5%) |
| **Phase** |  |
| Phase 2 | 2 (12.5%) |
| Phase 2–3 | 2 (12.5%) |
| Phase 3 | 12 (75.0%) |
| **Setting** |  |
| Metastatic | 16 (100.0%) |
| **Treatment line** |  |
| 1L | 12 (75.0%) |
| 2L | 2 (12.5%) |
| 2L+ | 2 (12.5%) |
| **Treatment type** |  |
| Chemo | 16 (100.0%) |

CRC, colorectal cancer; PDAC, pancreatic ductal adenocarcinoma; 1L, first-line; 2L, second-line; 2L+, second-line or beyond.

The KCS trial had no officially reported lead arm as it showed no significant difference in survival between arms. For this comparison, the arm with the higher observed median PFS was designated as the lead arm regardless of statistical significance. Results are reported both for the subset with officially reported lead arms (*n* = 7) and for the full set including this comparison (*n* = 8).

Among the seven comparisons with officially reported superiority of one arm, the model correctly identified the superior arm in 6 (86%; one-sided binomial test *p* = 0.06, 95% CI: 48%–100%). Extending the analysis to include one additional comparison in which no arm was officially designated as superior, using the arm with the higher observed mPFS as the reference, concordance remained high at 88% (7/8; *p* < 0.05, 95% CI: 53%–100%). The discordant prediction occurred in trial PMID14665611 (FOLFIRI vs. FOLFOX in CRC) for which the model predicted a mPFS difference of 0.32 months and ORR difference of 0.01 points with the FOLFIRI arm leading when the observed difference was 1.80 months and 0.14 points respectively, with the FOLFOX arm leading.

The model’s predictions aligned with observed outcomes even in cases where the mPFS difference between arms was modest. In trial KCS, where survival did not differ significantly between 5-FU and 5-FU + oxaliplatin, the model correctly identified the numerically superior arm. Across all 16 arms in the head-to-head subset, the model predicted mPFS with a mean absolute error of 0.78 months and ORR with a mean absolute error of 0.08 points, and the direction and relative magnitude of mPFS and ORR differences between arms were generally well captured, as shown in Figure 6.

**Figure 6:**
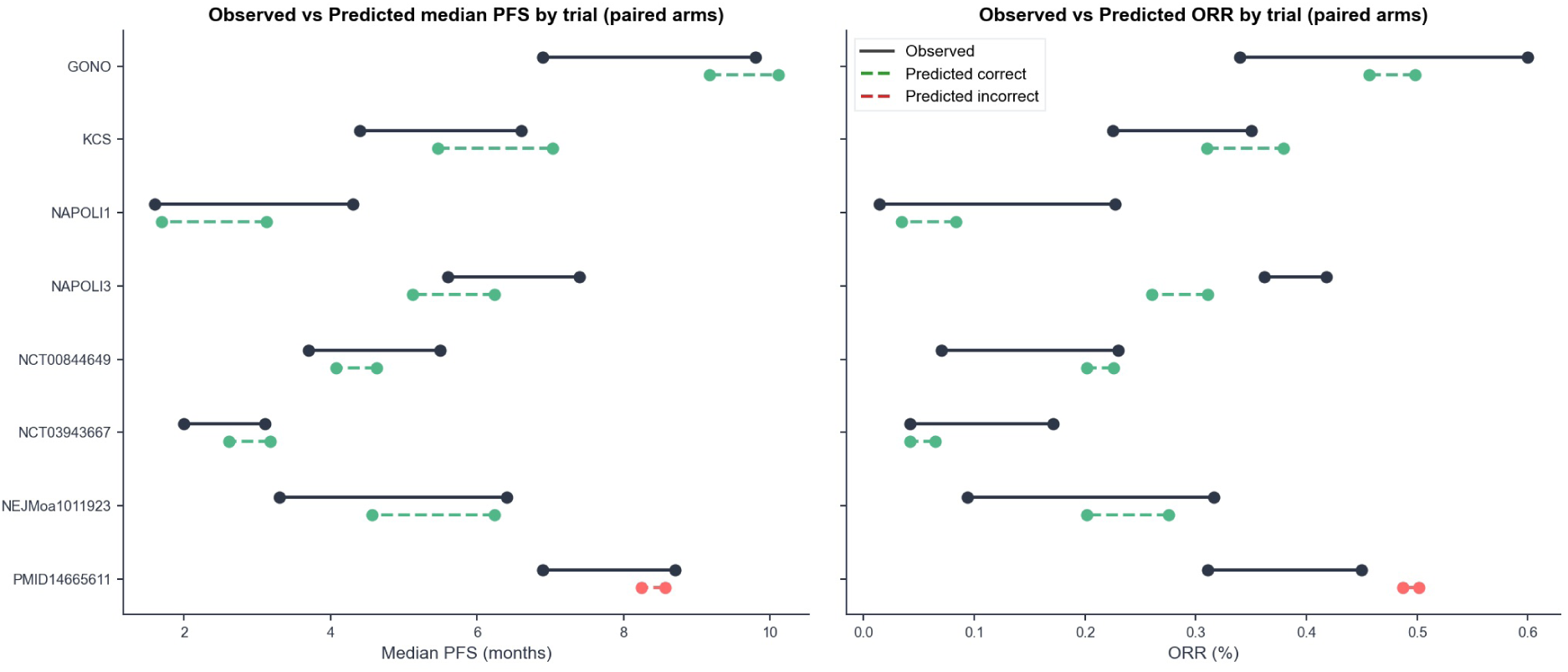
Concordance between predicted and observed mPFS and ORR across independent clinical trial comparisons. Each row represents a head-to-head trial comparison, with paired arms shown as connected points. Solid black lines indicate observed mPFS and ORR from published trial results; dashed lines indicate predictions. Green dashed lines denote comparisons where the model correctly identified the superior treatment arm; the red dashed line denotes the discordant prediction.

### Result 3: Prospective application to a novel RAS inhibitor – Daraxonrasib case study

To evaluate SCOPE in a prospective-style application, we extended the validation to Daraxonrasib (RMC-6236), a novel RAS(ON) multiselective inhibitor under evaluation in the RASolute clinical trial program for mPDAC. We simulated two head-to-head comparisons: Daraxonrasib versus FOLFIRINOX in second-line KRAS G12X mPDAC, and Daraxonrasib versus Gemcitabine plus nab-Paclitaxel (GnP) in first-line KRAS-mutant mPDAC. PDOs from the biobank that had been screened against Daraxonrasib were selected based on clinical characteristics matching the inclusion criteria of the different RASolute trials. Predicted outcomes were compared against preliminary phase 1 data for Daraxonrasib and published outcomes for the comparator regimens, as no phase 3 results were available at the time of analysis.

In the second-line setting, the platform predicted Daraxonrasib superiority, with a predicted mPFS of 8.9 versus 5.1 months and a predicted ORR of 29% versus 13% for FOLFIRINOX. Long-term followup data (15) in second-line KRAS G12X mPDAC reported a mPFS of 8.5 months (*n* = 26) and an ORR of 35%, while historical mFOLFIRINOX data (16) in second-line mPDAC in an unselected population report a mPFS of 3.9 months and an ORR of approximately 11% (*n*=104) (Figure 7A).

**Figure 7:**
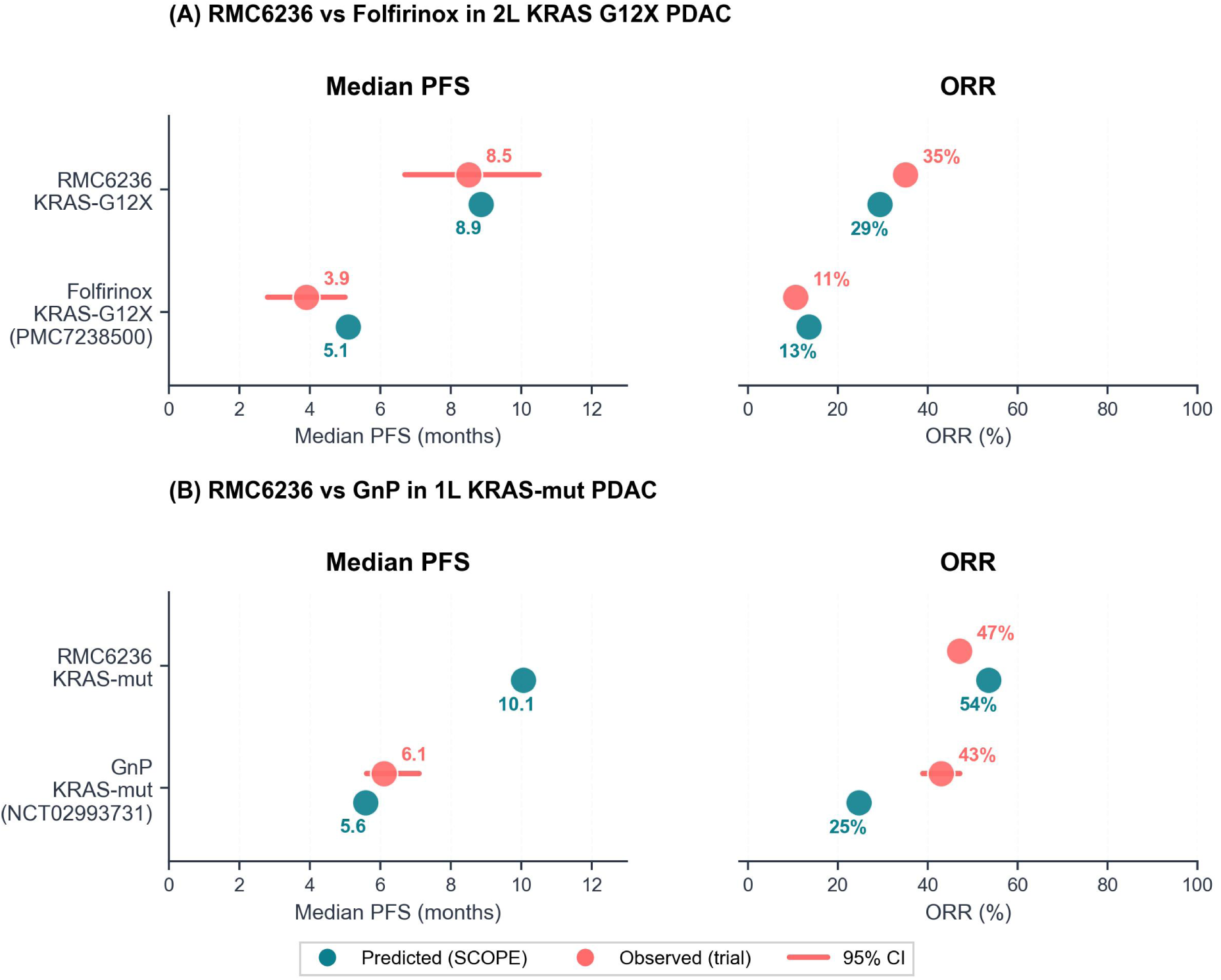
Prospective application of SCOPE to Daraxonrasib (RMC-6236) in metastatic PDAC. Predicted (teal) and observed (coral) ORR and median PFS for Daraxonrasib versus standard-of-care chemotherapy in two simulated trial scenarios. **(A)** Second-line setting (simulating RASolute-302), comparing Daraxonrasib to FOLFIRINOX in KRAS G12X mPDAC. Observed Daraxonrasib outcomes are from phase 1 long-term followup data (15). Observed FOLFIRINOX outcomes are from a retrospective series of mFOLFIRINOX in second-line mPDAC in an unselected population (16). **(B)** First-line setting (simulating RASolute-303), comparing Daraxonrasib to GnP in KRAS-mutant mPDAC. Observed Darax-onrasib ORR is from phase 1 data (15) in treatment-naïve KRAS-mutant patients; mPFS was not yet reported at data cutoff. Observed GnP outcomes are from a phase 3 trial in unselected first-line mPDAC (NCT02993731; (17)). Horizontal bars indicate 95% confidence intervals where available. GnP: gemcitabine plus nab-paclitaxel.

In the first-line setting, SCOPE again predicted Daraxonrasib as the superior arm, with a predicted median PFS of 10.1 versus 5.6 months and a predicted ORR of 54% versus 25% for GnP. Preliminary phase 1 data (15) from treatment-naïve RAS-mutant mPDAC patients reported an ORR of 47% for Daraxonrasib monotherapy (*n* = 38), consistent with the model’s prediction, while published GnP outcomes (17) in unselected mPDAC populations report a median PFS of 6.1 months and an ORR of 43% (*n*=559) (Figure 7B). In both settings, the predicted treatment effects were directionally consistent with the emerging clinical data. Notably, the comparator arm data available from the literature were derived from molecularly unselected populations, representing a conservative benchmark for the KRAS-selected trial populations simulated by SCOPE.

## DISCUSSION

This study demonstrates that integrating clinical prognostic scoring with patient-derived organoid drug screening enables calibrated, quantitative prediction of clinical trial outcomes in mCRC and mPDAC. Across 32 arms from 23 published trials, SCOPE achieved strong agreement with observed mPFS (*R*^2^ = 0.85, MAE < 1 month), correctly identified the superior treatment arm in 7 of 8 head-to-head comparisons, and produced predictions directionally consistent with emerging clinical data for Daraxonrasib, a novel pan-RAS inhibitor with no prior phase 3 data. To our knowledge, this is the first demonstration that PDO drug screening can be used to predict population-level treatment outcomes.

To contextualize model performance, we compared the agreement between predicted and observed mPFS against the concordance between two independently measured clinical end-points, observed ORR and observed mPFS, across the same trial arms. These two endpoints showed an *R*^2^ of 0.87, a level of agreement that can only be established by running the trial to completion and measuring both outcomes. SCOPE achieved 98% of this inter-endpoint concordance (*R*^2^ = 0.85) without access to any trial data, relying solely on pretrial information: baseline patient characteristics and in vitro drug sensitivity. This suggests that the platform captures a comparable share of arm-level outcome variance as an endpoint that requires full trial execution, positioning it as a tool to anticipate clinical benefit and inform trial design decisions before patient enrollment.

The ablation analysis confirmed that clinical and drug screening data provide complementary and non-redundant predictive information. The Clinical Score alone showed moderate concordance with observed outcomes but, because it is treatment-independent, produces identical predictions for all arms within a trial and therefore cannot perform treatment ranking. The Drug Screen Score provides the treatment-specific signal necessary to distinguish between regimens. The integrated model significantly outperformed either component in isolation, supporting the platform’s two-module architecture.

The single discordant prediction in head-to-head comparisons occurred in trial PMID14665611 (FOLFIRI vs. FOLFOX, first-line mCRC), where the model predicted a narrow margin of 0.32 months favoring FOLFIRI against an observed difference of 1.80 months favoring FOLFOX. This discrepancy is consistent with the well-documented comparability of these two regimens in multiple randomized trials (18; 19), which are considered interchangeable in current practice guidelines. The small predicted margin may itself be informative, reflecting genuine difficulty in separating two similarly effective treatments.

We acknowledge that the current validation set is enriched for comparisons between combination and monotherapy regimens, where the superior arm may appear clinically intuitive. A naïve heuristic (i.e. always predicting that the more intensive regimen will prevail) would achieve comparable concordance. However, SCOPE receives only a clinical prognostic score and organoid-derived drug sensitivity measurements, with no knowledge of treatment identity or regimen complexity. That an approach grounded in in vitro tumor biology converges with clinical intuition through an independent information pathway supports the biological validity of PDO-based drug screening. Unlike the naïve heuristic, SCOPE generalizes to settings where prior clinical intuition is unavailable, as demonstrated with the Daraxonrasib case study.

The Daraxonrasib case study illustrates the applicability of SCOPE to novel agents, including first-in-class therapies. In both the second-line and first-line mPDAC settings, predicted end-points were consistent with emerging data: the predicted mPFS of 8.9 months in the second-line setting closely matched the reported 8.5 months, and the predicted ORR of 54% in the first-line setting was consistent with the preliminary report of 47%. The comparator arm benchmarks used for these simulations were derived from molecularly unselected populations, representing a conservative reference for the KRAS-selected cohorts simulated by SCOPE. Beyond prediction, PDO-based platforms offer an additional advantage: the organoid panel can be inter-rogated to explore biological determinants of drug response, potentially informing biomarker development and patient selection strategies.

Several limitations should be considered. First, sample sizes are modest throughout the pipeline: the development cohort comprised 54 treatment lines, the validation set included 32 arms, and treatment ranking was assessed in 8 comparisons. Expanding both datasets is an active priority. Second, the validation set is heavily weighted towards cytotoxic chemotherapy (88% of arms). Benchmarking of the platform’s performance was limited by the existence of therapies that have been tested on the organoid platform and whose clinical trial results have been published. The performance of SCOPE on agents operating through or partially through immune-dependent pathways remains to be established and will be the subject of future development efforts. Furthermore, further development is required to model the efficacy of the anti-EGFR monoclonal antibodies cetuximab and panitumumab, whose efficacy depends on context-dependent EGFR pathway addiction, driven in part by ligand availability (e.g., AREG/EREG) and microenvironmental input that are not fully recapitulated in organoids.

Third, while substantial at 81 lines, the PDO biobank may not fully capture the molecular diversity of the trial populations. Fourth, the model exhibited regression-to-the-mean compression, tending to overpredict outcomes for poorly performing regimens and underpredict for highly effective ones. This likely reflects both the composition of the development cohort, consisting of heavily pretreated patients with limited treatment response, and the constraints of a linear prediction model. Given the limited size of the development cohort, we favored a linear model to preserve interpretability and reduce the risk of overfitting; non-linear approaches will be explored as more data become available. Fifth, ORR calibration was weaker in PDAC (*R*^2^ = 0.50) than in CRC (*R*^2^ = 0.85), possibly reflecting the narrower ORR range in pancreatic cancer trials. Finally, this study validates trial-level predictions only. SCOPE also generates patient-level predictions, for which external validation is ongoing in an independent cohort and will be reported separately. Clinical validation of patient-level predictions with SCOPE would pave the way towards the development of organoid-based precision medicine tools. These tools could ultimately help clinicians identify the optimal treatment regimens for their patients.

Expanding the development cohort and validation datasets, particularly with targeted therapy and ADC regimens, is the most immediate priority. We plan to improve trial population simulation by generating distributions of synthetic patient profiles rather than a single representative profile, which may reduce regression-to-the-mean compression. Non-linear modeling approaches will be explored once the development cohort is sufficiently powered. Extension to additional tumor types, including ovarian cancer and non-small cell lung cancer, is underway through disease-specific biobank development. Extending the framework to the prediction of overall survival is also a key objective, as overall survival remains the preferred primary end-point in registration trials. True prospective validation, in which predictions are registered before trial results become available, remains the definitive test and is being pursued as ongoing trials report primary endpoints.

SCOPE provides calibrated, quantitative predictions of clinical trial outcomes in mCRC and mPDAC by integrating clinical data with PDO drug screening. The platform correctly ranks competing treatments in the majority of head-to-head comparisons and extends to novel agents without established efficacy benchmarks. Although this initial validation is limited in scale, it establishes a proof of concept for organoid-informed trial outcome prediction and provides a foundation for prospective application in oncology drug development.

## Data Availability

The clinical trial validation dataset is provided in the supplementary material. Patient-level clinical and organoid drug screening data are not publicly available due to patient privacy considerations and proprietary restrictions. Requests for data access may be directed to the corresponding author and will be considered on a case-by-case basis subject to institutional and contractual agreements.

## Data sharing

The clinical trial validation dataset, including observed mPFS and ORR, is provided in the Supplementary Table 3. Patient-level clinical and organoid drug screening data are not publicly available due to patient privacy considerations and proprietary restrictions. Requests for data access may be directed to the corresponding author and will be considered on a case-by-case basis subject to institutional and contractual agreements.

