## Supplementary Tables for "SCOPE: Integrating Organoid Screening and Clinical Variables Through Machine Learning for Cancer Trial Outcome Prediction"

**Table 1A:** Patient demographics, disease characteristics, and molecular profile of the development cohort, reported per unique patient ($N$=52), stratified by cancer type.

| | All ($N$=52) | CRC ($N$=15) | PDAC ($N$=37) |
|---|---|---|---|
| **Patient demographics at diagnosis** | | | |
| Age, median [IQR] | 59.5 [50.5–67.0] | 55.0 [46.5–65.5] | 63.0 [53.0–67.0] |
| Sex | | | |
| Male | 27 (51.9%) | 7 (46.7%) | 20 (54.1%) |
| Female | 25 (48.1%) | 8 (53.3%) | 17 (45.9%) |
| BMI (kg/m$^2$), median [IQR] | 23.8 [21.0–25.0] | 24.0 [21.2–25.1] | 23.7 [20.8–25.0] |
| ECOG PS | | | |
| 0 | 0 (0.0%) | 0 (0.0%) | 0 (0.0%) |
| 1 | 26 (50.0%) | 13 (86.7%) | 13 (35.1%) |
| 2 | 17 (32.7%) | 2 (13.3%) | 15 (40.5%) |
| 3 | 4 (7.7%) | 0 (0.0%) | 4 (10.8%) |
| Missing | 5 (9.6%) | 0 (0.0%) | 5 (13.5%) |
| **Disease characteristics** | | | |
| Stage at diagnosis: metastatic | 52 (100.0%) | 15 (100.0%) | 37 (100.0%) |
| Prior surgery | 20 (38.5%) | 15 (100.0%) | 5 (13.5%) |
| Metastatic sites at diagnosis | | | |
| Liver | 39 (75.0%) | 10 (66.7%) | 29 (78.4%) |
| Peritoneal | 12 (23.1%) | 6 (40.0%) | 6 (16.2%) |
| Lung | 11 (21.2%) | 3 (20.0%) | 8 (21.6%) |
| Lymph nodes | 7 (13.5%) | 2 (13.3%) | 5 (13.5%) |
| No. of metastatic sites, median [range] | 1.0 [1.0–3.0] | 2.0 [1.0–3.0] | 1.0 [1.0–3.0] |
| **Molecular profile** | | | |
| MSI status: MSS | 50 (96.2%) | 15 (100.0%) | 35 (94.6%) |
| Missing | 2 (3.8%) | 0 (0.0%) | 2 (5.4%) |
| KRAS mutated | 43 (82.7%) | 12 (80.0%) | 31 (83.8%) |
| G12X | 36 (69.2%) | 6 (40.0%) | 30 (81.1%) |
| Other / Q61x | 7 (13.5%) | 6 (40.0%) | 1 (2.7%) |
| TP53 mutated | 38 (73.1%) | 10 (66.7%) | 28 (75.7%) |
| APC mutated | 9 (17.3%) | 9 (60.0%) | 0 (0.0%) |
| CDKN2A mutated | 8 (15.4%) | 0 (0.0%) | 8 (21.6%) |

Continuous variables are reported as median [IQR] or median [range]. Categorical variables are reported as $n$ (%). BMI, body mass index; CRC, colorectal cancer; ECOG PS, Eastern Cooperative Oncology Group performance status; IQR, interquartile range; MSI, microsatellite instability; MSS, microsatellite stable; PDAC, pancreatic ductal adenocarcinoma.

**Table 1B:** Organoid, treatment, and clinical outcome characteristics of the development cohort, reported per treatment line ($N$=54), as two patients contributed two distinct treatment lines each. Stratified by cancer type.

| | All ($N$=54) | CRC ($N$=15) | PDAC ($N$=39) |
|---|---|---|---|
| **PDO generation** | | | |
| PDO generation source | | | |
| Surgery | 1 (1.9%) | 1 (6.7%) | 0 (0.0%) |
| Core needle biopsy | 42 (77.8%) | 14 (93.3%) | 28 (71.8%) |
| Ascites | 10 (18.5%) | 0 (0.0%) | 10 (25.6%) |
| Pleural effusion | 1 (1.9%) | 0 (0.0%) | 1 (2.6%) |
| Sampling sites (biopsies and surgeries) | | | |
| Liver | 33 (61.1%) | 9 (60.0%) | 24 (61.5%) |
| Lung | 3 (5.6%) | 2 (13.3%) | 1 (2.6%) |
| Other | 7 (13.0%) | 4 (26.7%) | 3 (7.7%) |
| **Treatment characteristics** | | | |
| Metastatic lines before PDO sampling | | | |
| 0 | 1 (1.9%) | 0 (0.0%) | 1 (2.6%) |
| 1 | 8 (14.8%) | 2 (13.3%) | 6 (15.4%) |
| 2 | 17 (31.5%) | 3 (20.0%) | 14 (35.9%) |
| 3+ | 28 (51.9%) | 10 (66.7%) | 18 (46.2%) |
| Treatment regimen received after PDO sampling | | | |
| Chemotherapy | 43 (79.6%) | 6 (40.0%) | 37 (94.9%) |
| Targeted therapy | 7 (13.0%) | 5 (33.3%) | 2 (5.1%) |
| Chemotherapy + targeted therapy | 4 (7.4%) | 4 (26.7%) | 0 (0.0%) |
| **Clinical outcomes after PDO sampling** | | | |
| PFS (months), median [IQR] | 2.0 [1.6–2.6] | 2.0 [1.7–3.2] | 2.1 [1.6–2.6] |
| Responder (SD or better) | 4 (7.4%) | 1 (6.7%) | 3 (7.7%) |

Continuous variables are reported as median [IQR] or median [range]. Categorical variables are reported as $n$ (%). CRC, colorectal cancer; IQR, interquartile range; PDAC, pancreatic ductal adenocarcinoma; PDO, patient-derived organoid; PFS, progression-free survival; SD, stable disease.

**Table 2A:** Baseline characteristics of the PDO biobank: patient demographics, disease characteristics, and molecular profile ($N$=78), stratified by cancer type.

| | **All ($N$=78)** | **CRC ($N$=42)** | **PDAC ($N$=36)** |
|---|---|---|---|
| **Demographics** | | | |
| Age at diagnosis, median [IQR] | 56.0 [48.0–67.5] | 55.0 [48.0–66.0] | 58.5 [51.8–67.5] |
| Missing | 2 (2.6%) | 2 (4.8%) | 0 (0.0%) |
| Sex | | | |
| Male | 41 (52.6%) | 22 (52.4%) | 19 (52.8%) |
| Female | 37 (47.4%) | 20 (47.6%) | 17 (47.2%) |
| ECOG PS at diagnosis | | | |
| 1 | 50 (64.1%) | 32 (76.2%) | 18 (50.0%) |
| 2 | 17 (21.8%) | 7 (16.7%) | 10 (27.8%) |
| 3 | 4 (5.1%) | 0 (0.0%) | 4 (11.1%) |
| 4 | 1 (1.3%) | 0 (0.0%) | 1 (2.8%) |
| Missing | 6 (7.7%) | 3 (7.1%) | 3 (8.3%) |
| **Disease characteristics** | | | |
| Stage at diagnosis | | | |
| Metastatic | 71 (91.0%) | 36 (85.7%) | 35 (97.2%) |
| Non-metastatic | 5 (6.4%) | 4 (9.5%) | 1 (2.8%) |
| Missing | 2 (2.6%) | 2 (4.8%) | 0 (0.0%) |
| Primary surgery | 39 (50.0%) | 34 (81.0%) | 5 (13.9%) |
| Metastatic site | | | |
| Liver | 59 (75.6%) | 30 (71.4%) | 29 (80.6%) |
| Lymph nodes | 19 (24.4%) | 11 (26.2%) | 8 (22.2%) |
| Lung | 18 (23.1%) | 9 (21.4%) | 9 (25.0%) |
| Peritoneal | 16 (20.5%) | 10 (23.8%) | 6 (16.7%) |
| Bone | 4 (5.1%) | 2 (4.8%) | 2 (5.6%) |
| Other | 7 (9.0%) | 6 (14.3%) | 1 (2.8%) |
| **Molecular profile** | | | |
| MSI status | | | |
| MSS | 69 (88.5%) | 36 (85.7%) | 33 (91.7%) |
| MSI | 3 (3.8%) | 2 (4.8%) | 1 (2.8%) |
| Missing | 6 (7.7%) | 4 (9.5%) | 2 (5.6%) |
| KRAS mutated | 55 (70.5%) | 28 (66.7%) | 27 (75.0%) |
| Any G12 variant | 43 (55.1%) | 17 (40.5%) | 26 (72.2%) |
| Other KRAS variant | 12 (15.4%) | 11 (26.2%) | 1 (2.8%) |

Continuous variables are reported as median [IQR]. Categorical variables are reported as $n$ (%). CRC, colorectal cancer; ECOG PS, Eastern Cooperative Oncology Group performance status; IQR, interquartile range; MSI, microsatellite instability; MSS, microsatellite stable; PDAC, pancreatic ductal adenocarcinoma; PDO, patient-derived organoid.

**Table 2B:** PDO line, treatment history, drug screening, and drug combination characteristics, reported per PDO line ($N$=81), as three patients contributed two PDO lines each. Stratified by cancer type.

| | **All ($N$=81)** | **CRC ($N$=44)** | **PDAC ($N$=37)** |
|---|---|---|---|
| **PDO line characteristics** | | | |
| Tissue source | | | |
| Biopsy | 60 (74.1%) | 35 (79.5%) | 25 (67.6%) |
| Ascites | 10 (12.3%) | 0 (0.0%) | 10 (27.0%) |
| Surgery | 11 (13.6%) | 9 (20.5%) | 2 (5.4%) |
| Biopsy or surgery site | | | |
| Liver | 51 (63.0%) | 31 (70.5%) | 20 (54.1%) |
| Peritoneum | 6 (7.4%) | 5 (11.4%) | 1 (2.7%) |
| Lung | 2 (2.5%) | 1 (2.3%) | 1 (2.7%) |
| Lymph node | 1 (1.2%) | 1 (2.3%) | 0 (0.0%) |
| Primary tumor | 2 (2.5%) | 0 (0.0%) | 2 (5.4%) |
| Other | 7 (8.6%) | 6 (13.6%) | 1 (2.7%) |
| Missing | 2 (2.5%) | 0 (0.0%) | 2 (5.4%) |
| **Treatment history** | | | |
| Metastatic lines before sampling | | | |
| 0 | 10 (12.3%) | 5 (11.4%) | 5 (13.5%) |
| 1 | 11 (13.6%) | 3 (6.8%) | 8 (21.6%) |
| 2 | 26 (32.1%) | 12 (27.3%) | 14 (37.8%) |
| 3+ | 34 (42.0%) | 24 (54.5%) | 10 (27.0%) |
| **Drug screening** | | | |
| Total drug–PDO pairs | 1295 | 698 | 597 |
| Single agents tested | 9 | 9 | 9 |
| Combinations tested | 4 | 4 | 4 |
| Drugs per PDO line, median [IQR] | 11.0 [6.0–11.0] | 11.0 [5.0–11.0] | 9.0 [6.0–11.0] |
| $EC_{50}$ (plate-norm), median [IQR] | 6.6 [5.6–7.8] | 6.6 [5.7–7.8] | 6.5 [5.6–7.9] |
| Max response (plate-norm), median [IQR] | 0.8 [0.7–0.9] | 0.8 [0.7–0.9] | 0.8 [0.7–0.9] |
| **Drugs screened by class** | | | |
| *Cytotoxic chemotherapy* | | | |
| 5-FU | 63 lines | 35 lines | 28 lines |
| Oxaliplatin | 57 lines | 31 lines | 26 lines |
| SN-38 | 67 lines | 36 lines | 31 lines |
| Gemcitabine | 57 lines | 32 lines | 25 lines |
| Paclitaxel | 64 lines | 32 lines | 32 lines |
| Trifluridine/tipiracil | 46 lines | 28 lines | 18 lines |
| *Targeted therapy – KRAS inhibitors* | | | |
| RMC-6236 | 72 lines | 39 lines | 33 lines |
| *Antibody–drug conjugates* | | | |
| Sacituzumab govitecan | 15 lines | 8 lines | 7 lines |
| Tusamitamab ravtansine | 6 lines | 3 lines | 3 lines |
| **Drug combinations** | | | |
| PDO lines screened with combos | 75 | 40 | 35 |
| 5-FU + Oxaliplatin + SN-38 | 72 lines | 39 lines | 33 lines |
| 5-FU + Oxaliplatin | 73 lines | 40 lines | 33 lines |
| Gemcitabine + Paclitaxel | 52 lines | 29 lines | 23 lines |
| 5-FU + SN-38 | 51 lines | 29 lines | 22 lines |

Continuous variables are reported as median [IQR]. Categorical variables are reported as $n$ (%). $EC_{50}$ and max response values are reported across all drug–PDO pairs after plate-level normalization. CRC, colorectal cancer; $EC_{50}$, half-maximal effective concentration; IQR, interquartile range; PDAC, pancreatic ductal adenocarcinoma; PDO, patient-derived organoid.

**Table 3:** SCOPE calibration performance metrics for median PFS and ORR predictions across the clinical trial validation dataset, stratified by cancer type.

**A) mPFS prediction metrics**

| | **Pearson** $r$ | **Pearson** $p$ | $R^2$ | **Spearman** $\rho$ | **Spearman** $p$ | MAE | RMSE | ±**1M** | ±**2M** | ±**3M** |
|---|---|---|---|---|---|---|---|---|---|---|
| CRC (14) | 0.91 | <0.001 | 0.82 | 0.80 | <0.001 | 0.94 | 1.14 | 0.57 | 0.93 | 1.00 |
| PDAC (17) | 0.89 | <0.001 | 0.80 | 0.84 | <0.001 | 0.73 | 0.87 | 0.65 | 1.00 | 1.00 |
| All (31) | 0.92 | <0.001 | 0.85 | 0.91 | <0.001 | 0.82 | 1.00 | 0.61 | 0.97 | 1.00 |

**B) ORR prediction metrics**

| | **Pearson** $r$ | **Pearson** $p$ | $R^2$ | **Spearman** $\rho$ | **Spearman** $p$ | **MAE (%)** | **RMSE (%)** | ±**10%** | ±**20%** | ±**30%** |
|---|---|---|---|---|---|---|---|---|---|---|
| CRC (15) | 0.92 | <0.001 | 0.85 | 0.84 | <0.001 | 6.27 | 7.92 | 0.80 | 1.00 | 1.00 |
| PDAC (17) | 0.71 | 0.002 | 0.50 | 0.82 | <0.001 | 8.26 | 9.91 | 0.47 | 0.94 | 1.00 |
| All (32) | 0.84 | <0.001 | 0.71 | 0.86 | <0.001 | 7.32 | 9.03 | 0.62 | 0.97 | 1.00 |

MAE, mean absolute error; mPFS, median progression-free survival; ORR, objective response rate; $R^2$, coefficient of determination; RMSE, root mean square error.

**Table 4:** Clinical trial validation dataset. Each row represents a treatment arm from a published clinical trial in metastatic CRC or PDAC.

| Trial ID | Dis. | Line | Phase | Year | ECOG | Age | Subgroup | Arm Drug | Type | mPFS (95% CI) | ORR (95% CI) |
|---|---|---|---|---|---|---|---|---|---|---|---|
| GONO [1] | CRC | 1L | 3 | 2001 | ≤1 | 60 | | Folfiri | Chemo | 6.90 | 34.00 |
| GONO [1] | CRC | 1L | 3 | 2001 | ≤1 | 60 | | Folfirinox | Chemo | 9.80 | 60.00 |
| KCS [2] | CRC | 1L | 2 | 2006 | ≤2 | 74 | | 5FU | Chemo | 4.40 | 22.50 (9.6–35.4) |
| KCS [2] | CRC | 1L | 2 | 2006 | ≤2 | 74 | | Folfox | Chemo | 6.60 | 35.00 (20.2–49.8) |
| NCT00069108 | CRC | 2L | 3 | 2003 | ≤2 | 60 | | Folfox | Chemo | 5.52 (4.77–5.98) | 20.13 |
| NCT00154102 | CRC | 1L | 3 | 2004 | ≤1 | 60 | EGFR Expr. | Folfiri | Chemo | 8.00 (7.6–9.0) | 38.70 (34.8–42.8) |
| NCT00339183 | CRC | 2L | 3 | 2006 | ≤2 | 61 | KRAS Mut. | Folfiri | Chemo | 4.90 (3.6–5.6) | 13.92 (9.78–19.00) |
| NCT00364013 [3] | CRC | 1L | 3 | 2006 | ≤2 | 61 | KRAS WT | Folfox | Chemo | 8.60 (7.5–9.5) | 48.00 (42.0–53.1) |
| NCT00364013 [3] | CRC | 1L | 3 | 2006 | ≤2 | 61 | KRAS Mut. | Folfox | Chemo | 9.20 (8.1–9.9) | 41.00 (34.1–47.7) |
| NCT01631552 [4] | CRC | 2L+ | 1–2 | 2012 | ≤1 | 60 | | Sac. Gov. | ADC | 3.90 (1.9–5.6) | 3.20 (0.1–16.7) |
| NCT01955837 | CRC | 3L+ | 3 | 2012 | ≤1 | 58 | | TFD/TPI | Chemo | 2.00 (1.9–2.8) | 1.10 (0.2–3.3) |
| NCT02187848 [5] | CRC | 1L+ | 1 | 2014 | ≤1 | 59 | | Tus. Rav. | ADC | – | 11.00 |
| PMID14665611 [6] | CRC | 1L | 3 | 2000 | ≤1 | 65 | | Folfiri | Chemo | 6.90 | 31.10 |
| PMID14665611 [6] | CRC | 1L | 3 | 2000 | ≤1 | 65 | | Folfox | Chemo | 8.70 | 45.00 |
| PMID22855138 [7] | CRC | 1L | 2 | 2002 | ≤2 | 61 | | Folfox | Chemo | 8.70 | 36.30 |
| CALGB80303 [8] | PDAC | 1L | 3 | 2004 | ≤2 | 64 | | Gemcitabine | Chemo | 2.90 (2.4–3.7) | 10.00 |
| GOIRC [9] | PDAC | 1L | 2 | 2004 | ≤2 | 64 | | Gemcitabine | Chemo | 3.50 (0.5–16.2) | 8.00 (0.5–16) |
| NAPOLI1 [10] | PDAC | 2L | 3 | 2012 | ≤2 | 62 | | Folfiri | Chemo | 4.30 (3.1–5.7) | 22.70 |
| NAPOLI1 [10] | PDAC | 2L | 3 | 2012 | ≤2 | 62 | | 5FU + leucovorin | Chemo | 1.60 (1.4–2.6) | 1.40 |
| NAPOLI3 [11] | PDAC | 1L | 3 | 2020 | ≤1 | 63 | | Folfirinox | Chemo | 7.40 (6.0–7.7) | 41.80 (36.8–46.9) |
| NAPOLI3 [11] | PDAC | 1L | 3 | 2020 | ≤1 | 63 | | GemPax | Chemo | 5.60 (5.3–5.8) | 36.20 (31.4–41.2) |
| NCT00844649 | PDAC | 1L | 3 | 2009 | ≤1 | 62 | | GemPax | Chemo | 5.50 (4.47–5.95) | 23.00 (19.1–27.2) |
| NCT00844649 | PDAC | 1L | 3 | 2009 | ≤1 | 62 | | Gemcitabine | Chemo | 3.70 (3.61–4.04) | 7.00 (5.0–10.1) |
| NCT01631552 [4] | PDAC | 2L+ | 1–2 | 2012 | ≤1 | 60 | | Sac. Gov. | ADC | 2.00 (1.1–3.5) | 0.00 (0–20.6) |
| NCT02993731 [12] | PDAC | 1L | 3 | 2017 | ≤1 | 64 | | GemPax | Chemo | 6.10 (5.6–7.1) | 42.90 (38.8–47.2) |
| NCT03943667 [13] | PDAC | 2L+ | 3 | 2019 | ≤2 | 64 | | GemPax | Chemo | 3.10 (2.2–4.3) | 17.10 (11.3–24.4) |
| NCT03943667 [13] | PDAC | 2L+ | 3 | 2019 | ≤2 | 64 | | Gemcitabine | Chemo | 2.00 (1.9–2.3) | 4.20 (0.9–11.9) |
| NCT06625320 [14] | PDAC | 2L | 1 | 2024 | ≤1 | 64 | G12X | RMC-6236 | TT | 8.50 (6.7–10.5) | 35.00 |
| NEJMoa1011923 [15] | PDAC | 1L | 2–3 | 2005 | ≤1 | 61 | | Folfirinox | Chemo | 6.40 (5.5–7.2) | 31.60 |
| NEJMoa1011923 [15] | PDAC | 1L | 2–3 | 2005 | ≤1 | 61 | | Gemcitabine | Chemo | 3.30 (2.2–3.6) | 9.40 |
| PMID17452677 [16] | PDAC | 1L | 3 | 2001 | ≤2 | 60 | | Gemcitabine | Chemo | 3.55 | 8.00 |
| S0205 [17] | PDAC | 1L | 3 | 2004 | ≤2 | 64 | | Gemcitabine | Chemo | 3.00 | 14.00 |

Trial NCT02187848 has no published mPFS. Year designates the year of enrolment start. ADC: antibody–drug conjugate; Chemo: chemotherapy; CI: confidence interval; CRC: colorectal cancer; Dis: Disease; ECOG: Eastern Cooperative Oncology Group; EGFR: epidermal growth factor receptor; Expr.: expressing; G12X: KRAS G12X mutant; Mut.: mutant; ORR: objective response rate; PDAC: pancreatic ductal adenocarcinoma; mPFS: median progression-free survival; Sac. Gov.: sacituzumab govitecan; TFD/TPI: trifluridine/tipiracil; TT: targeted therapy; Tus. Rav.: tusamitamab ravtansine; WT: wild-type.

**Table 5:** Head-to-head treatment arm ranking results. For each trial with two or more evaluable arms, the observed and predicted leading treatment arm are compared. Observed and predicted mPFS and ORR differences are reported alongside log-rank $P$-values and hazard ratios where available.

| **Trial ID** | **Disease** | **Arms / treatments** | **Obs. mPFS Δ** | **Pred. mPFS Δ** | **Obs. ORR Δ** | **Pred. ORR Δ** | **Obs. lead arm** | **Pred. lead arm** | **Off. lead arm** | **Obs. log-rank $P$** | **Obs. HR (95% CI)** |
|---|---|---|---|---|---|---|---|---|---|---|---|
| NCT03943667 | PDAC | GemPax vs Gemcitabine | 1.10 | 0.56 | 0.13 | 0.02 | GemPax | GemPax | GemPax | $<$0.01 | 0.64 (0.47–0.89) |
| KCS | CRC | 5FU vs. Folfox | 2.20 | 1.65 | 0.12 | 0.07 | Folfox | Folfox | - | 0.34 | 1.33 (0.74–2.37) |
| GONO | CRC | Folfiri vs Folfirinox | 2.90 | 0.77 | 0.26 | 0.04 | Folfirinox | Folfirinox | Folfirinox | $<$0.001 | 0.63 |
| PMID14665611 | CRC | Folfiri vs Folfox | 1.80 | 0.32 | 0.14 | 0.01 | Folfox | Folfiri | Folfox | $<$0.005 | – |
| NEJMoa1011923 | PDAC | Folfirinox vs Gemcitabine | 3.10 | 1.65 | 0.22 | 0.07 | Folfirinox | Folfirinox | Folfirinox | $<$0.001 | 0.47 (0.37–0.59) |
| NAPOLI3 | PDAC | Folfirinox vs GemPax | 1.80 | 1.10 | 0.06 | 0.05 | Folfirinox | Folfirinox | Folfirinox | $<$0.0001 | 0.69 (0.58–0.83) |
| NCT00844649 | PDAC | GemPax vs Gemcitabine | 1.80 | 0.56 | 0.16 | 0.02 | GemPax | GemPax | GemPax | $<$0.0001 | 0.69 (0.58–0.82) |
| NAPOLI1 | PDAC | Folfiri vs 5FU + leucovorin | 2.70 | 1.44 | 0.21 | 0.05 | Folfiri | Folfiri | Folfiri | $<$0.0001 | – |

CI, confidence interval; CRC, colorectal cancer; HR, hazard ratio; –, not available; Obs., observed; Off., official; ORR, objective response rate; PDAC, pancreatic ductal adenocarcinoma; PFS, progression-free survival; Pred., predicted. Obs. mPFS Δ is the absolute difference of mPFS between arms. Pred. mPFS Δ is the absolute difference in SCOPE–predicted mPFS between arms.
